# Safety and Biological Activity of Intravitreal OGX110, a CXCR3 Agonist, in Persistent Neovascular Age-Related Macular Degeneration: A Phase I Dose-Escalation Study

**DOI:** 10.64898/2026.05.21.26353430

**Authors:** Alan Wells, David Boyer, Roger A. Goldberg, Thomas Hohman, Raj Maturi, Sunil Patel

## Abstract

**Purpose:** To evaluate the safety and exploratory outcomes of a single intravitreal injection of OGX110, a peptide agonist of CXCR3, in eyes with persistent fluid secondary to neovascular age-related macular degeneration (nAMD) despite ongoing anti-vascular endothelial growth factor (anti-VEGF) therapy.

**Methods:** This prospective, open-label, sequential dose-escalation phase I study (NCT05904691) enrolled subjects receiving standard-of-care intravitreal anti-VEGF therapy. Subjects received a single intravitreal injection of OGX110 at 0.5 mg, 1.0 mg, or 2.0 mg (n=3 per cohort), 7 to 14 days after the anti-VEGF injection.

**Results:** All nine enrolled subjects completed follow-up through day 56. Two subjects (22%) experienced at least 1 adverse event (AE); all were mild and unrelated to study treatment. Exploratory analyses showed a BCVA change of +1.4 letters following anti-VEGF injection and +4.4 letters from OGX110 baseline to 4 weeks (P < 0.05). Six of 9 subjects gained at least 3 ETDRS letters after OGX110. Anatomic responses were heterogeneous. Four eyes showed a reduction in CRT after anti-VEGF injection that was maintained after OGX110 administration. One additional eye demonstrated a substantial reduction in CRT after OGX110 despite minimal response to anti-VEGF treatment.

**Conclusions:** A single intravitreal injection of OGX110 was well tolerated. Exploratory functional and anatomic findings suggest biologic activity; interpretation is limited by small sample size, open-label design, absence of a concurrent control group, and inter-subject heterogeneity. These results support further study in a controlled trial.

**Translational Relevance:** OGX110 represents a mechanistically distinct investigational approach for nAMD that may warrant further evaluation in eyes with persistent.

## Introduction

Age-related macular degeneration (AMD) is the leading cause of visual impairment in adults over 50 [1-4]. A substantial fraction of subjects with this progressive disease develops choroidal neovascularization (nAMD) with the formation of new blood vessels that are immature and leak. Excessive extravascular fluid including hemorrhage and other inflammatory matrix deposits creates a feedback loop in which the continued production of inflammatory cytokines and growth factors not only increases the intraretinal, subretinal, and sub-retinal pigment epithelium (RPE) fluid, but also eventually leads to scarring with atrophic retinal cell death and blindness. This sequence of events, while presenting unique aspects, does follow the pattern of a festering wound undergoing chronic sterile inflammation [5-7].

The incipient insult that starts this non-resolving wound is still ill-defined, and likely multifactorial [8]. Still, a key driver in the early vision loss is excessive levels of vascular endothelial growth factor (VEGF). This factor leads to de-cohesion of the endothelial cells in the retinal capillaries promoting intravascular fluid leakage into the extravascular spaces and enabling neo-angiogenesis. To counter these deleterious effects of VEGF, VEGF-sequestering therapies (both anti-VEGF antibodies and VEGF traps) have been deployed to beneficial effect [9]. Two-thirds of newly diagnosed subjects, in controlled clinical trials, show demonstrable visual benefit (at least one line gain) from intravitreal injections of anti-VEGF agents, improving or stabilizing eyesight over the first year. However, the effectiveness of these treatments can wane over several years of treatment, with a difficult-to-sustain treatment burden that can lead can lead to persistent disease activity, subsequent fibrosis and atrophy, and vision loss over time [10-13].

The limited value of longer-term anti-VEGF therapies and alternative competitive inhibitors could be due to nAMD being a wound chronically in the tissue replacement phase that does not progress to resolution [8]. Wounds usually transition to the resolution phase after sensing tissue closure, but in the absence of this in nAMD, due to the physical anatomy of the ocular cavity being an ‘unclosed’ space and/or the continued sterile inflammatory milieu driven by the drusen [5-7], the tissue does not recognize the drivers of resolution [14]. As the nAMD wound does not resolve, the drivers of angiogenesis and scarring remain, and become dominant when the competitive inhibitors fall below effective concentrations. The tissue replacement phase in healing wounds is marked by an inflammatory milieu that drives both neo-angiogenesis and excessive matrix deposition [14, 15]. During resolution, up to 90% of the new vessels involute, the matrix is resorbed and remodeled, and the inflammation suppressed. Wounds that fail to transition to resolution remain exudative, and despite continual anti-VEGF therapy, may continue to hypertrophic scarring and atrophy.

Combating this persistent wound healing response may require an agent that can trigger resolution and thus drive nAMD into remission. In a variety of tissues, this is accomplished physiologically by chemokines that activate the C-X-C motif chemokine receptor 3 (CXCR3) seven transmembrane G-protein coupled receptor [6, 16-18]; these are acute phase reactants whose expression is increased multifold upon contact inhibition of endothelial and epithelial cells in the wound [14]. In the absence either the receptor or its cognate chemokines (CXCL10 and CXCL11) the wound remains in the sterile inflammatory stage and progresses over months to hypertrophic and hypervascular scarring. In the reverse, scarring can be limited in preclinical models such as IPF and renal fibrosis by administration of pharmacologic levels of these chemokines [18, 19]. nAMD presents an opportunity to apply similar treatment, in that the CXCR3 receptor is present in this disease and the chemokines C-X-C motif chemokine ligand 10 (CXCL10) and C-X-C motif chemokine ligand 11 (CXCL11) do not evince the multifold increased expression noted during physiologic wound resolution [20].

The present first-in-human phase I study was designed primarily to assess the safety and tolerability in humans of a novel peptide agonist of CXCR3 that was derived from the receptor-binding sequence of CXCL10 [21]. Provided in this report is the single-ascending dose portion of this study. Subjects with persistent nAMD activity despite prior anti-VEGF therapy received a single intravitreal injection of OGX110 (also formally as OCU-10-C-110) across 3 ascending dose levels. Functional and anatomic outcomes were assessed as exploratory end points.

## Methods

### Study Design

This was a prospective, open-label, sequential dose-escalation phase I study evaluating the safety and tolerability of a single intravitreal injection of OGX110 in subjects with active nvAMD (ClinicalTrials.gov identifier: NCT05904691).

Subjects were enrolled sequentially into 3 dose cohorts: 0.5 mg, 1.0 mg, and 2.0 mg (3 subjects per cohort). OGX110 was administered as a single 50-uL intravitreal injection 7 to 14 days after the most recent standard-of-care anti-VEGF injection. Subjects returned on day 1, week 2, week 4, and week 8 (day 56) after OGX110 administration. Dose escalation proceeded only after protocol-specified safety review of the preceding cohort by the safety review committee (Figure 1).

**Figure 1.**
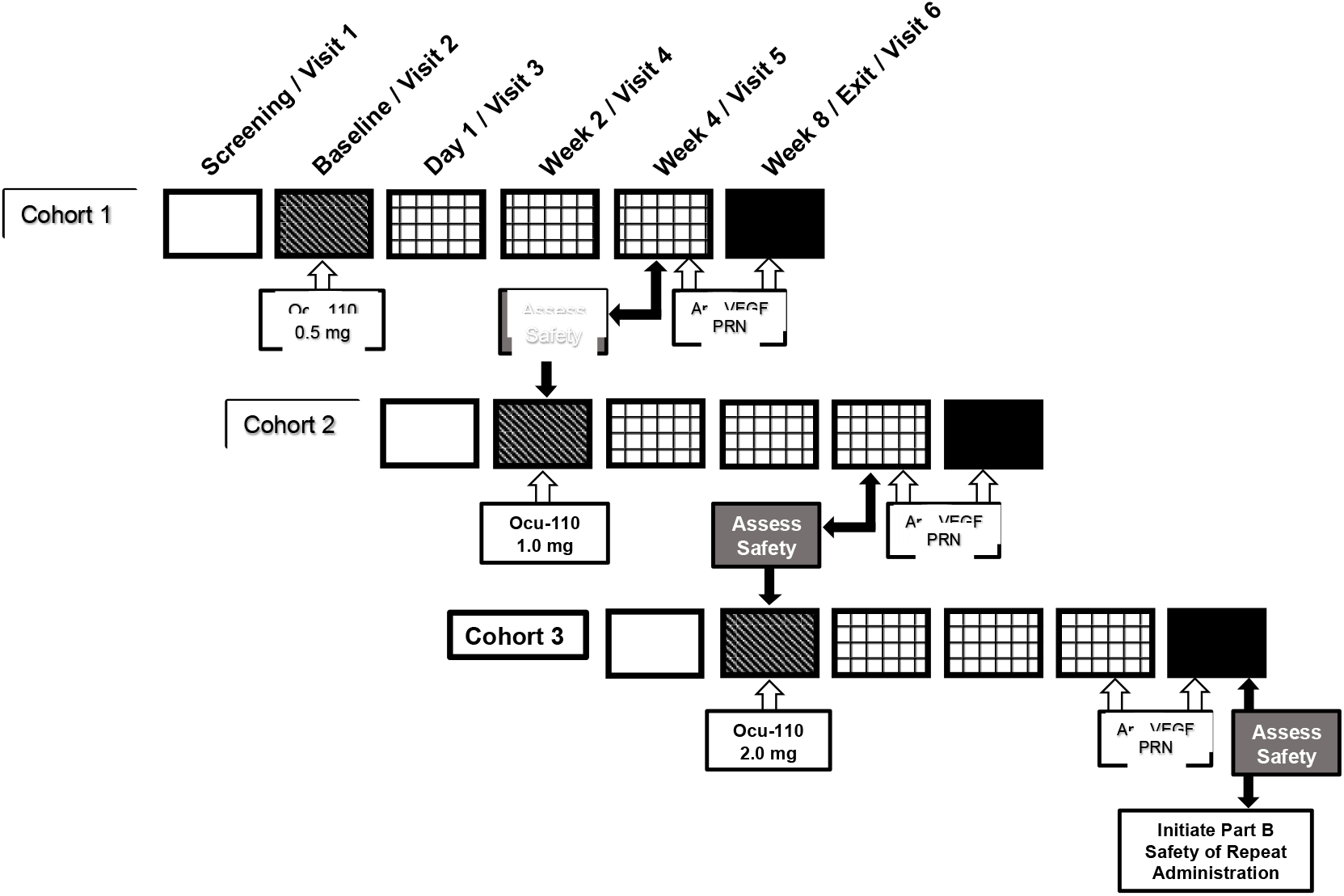
Schematic of study with cohorts and visit and safety check schedules

Rescue anti-VEGF therapy was permitted at investigator discretion at week 4 and week 8, or earlier if clinically indicated.

### Study Population

Eligible for enrollment were individuals aged 50 years or older with angiographically documented active choroidal neovascularization secondary to AMD with evidence of exudative activity documented within 10 weeks before enrollment. All study eyes had received an intravitreal anti-VEGF agent approved in the United States 7 to 14 days before OGX110 administration. BCVA in the study eye was required to be between 24 and 78 ETDRS (Early Treatment of Diabetic Retinopathy Study) letters at screening and baseline. Full inclusion and exclusion criteria are provided in Supplementary Tables 1 and 2.

The protocol was approved by a central institutional review board, and all individuals provided written informed consent. The study adhered to the tenets of the Declaration of Helsinki and in compliance with International Conference on Harmonisation guidelines of Good Clinical Practice and applicable national and local ethical and legal requirements.

### Study Drug Administration

OGX110 was supplied as a sterile aqueous solution formulated with standard excipients for intravitreal injection in single-use vials and stored at 2 C to 8 C until use. Each injection was administered in a volume of 50 uL using standard intravitreal injection technique.

### Study Outcomes

The primary objective was safety and tolerability. Safety assessments included adverse events, ophthalmic examinations, intraocular pressure measurement, BCVA, and SD-OCT. Additional baseline and exit assessments included fluorescein angiography and fundus photography. The visit schedule is summarized in Supplementary Table 3. All data was collected and validated by the third party CRO. All analyses of the data were performed *a posteriori* by the investigative team using standard statistical formula; the BCVA changes were calculated using the Student’s t-Test, paired (for intra-patient changes), one-sided.

## Results

### Subjects and Safety

Twelve subjects were screened, and 9 were enrolled and treated. Three screen failures occurred because fluorescein angiography did not demonstrate sufficient leakage to confirm active disease. All 9 treated subjects completed follow-up through day 56, and no subject required rescue therapy during the study period (all enrolled subjects completed the flow chart in Figure 1).

Baseline demographic and ocular characteristics are summarized in Table 1. Subjects ranged in age from 58 to 87 years. Study eyes had a history of nAMD ranging from 3 months to 9 years and had been treated with several commonly used anti-VEGF agents, including bevacizumab, aflibercept, ranibizumab, and faricimab. Baseline BCVA ranged from 30 to 70 ETDRS letters, and baseline CRT ranged from 196 to 557 um.

**Table 1.** Patient Demographics.

| **Patient #** | **Sex** | **Age (years)** | **nvAMD yr/mn** | **Anti-VEGF** | **BCVA (ETDRS)** | **CRT (microns)** | **Cohort** |
| --- | --- | --- | --- | --- | --- | --- | --- |
| 1 | M | 71-75 | 1yr 11mn | Ranibizumab-eqrn | 70 | 250 | Low/0.5mg |
| 2 | M | 56-60 | 0y 3mn | Bevacizumab | 46 | 557 | Low/0.5mg |
| 3 | F | 71-75 | 1yr 9mn | Ranibizumab-eqrn | 60 | 374 | Low/0.5mg |
| 4 | M | 86-90 | 0yr 3mn | Ranibizumab-eqrn | 70 | 330 | Mid/1.0mg |
| 5 | M | 76-80 | 5yr 3mn | Faricimab | 57 | 196 | Mid/1.0mg |
| 6 | F | 76-80 | 0yr 3mn | Aflibercept | 55 | 428 | Mid/1.0mg |
| 7 | F | 76-80 | 0yr 7mn | Ranibizumab-eqrn | 30 | 380 | High/2.0mg |
| 8 | F | 81-85 | 9yr 0mn | Aflibercept | 68 | 261 | High/2.0mg |
| 9 | F | 71-75 | 3yr 9mn | Faricimab | 65 | 199 | High/2.0mg |

M = male; F = Female
nvAMD = Neovascular age-related macular degeneration – duration since diagnosis in the study eye.
BCVA = Best correct visual acuity in Early Treatment of Diabetic Retinopathy Study letters.

OGX110 was generally well tolerated across all 3 dose levels. Two of 9 subjects experienced at least 1 AE. All reported AEs were mild and judged unrelated to study treatment: one subject in the mid-dose cohort was noted to have a grade 1 vitreous hemorrhage at day 14 that had resolved by day 28, and one subject in the high-dose cohort had an epiretinal membrane noted at screening and again at day 1 and day 56, and also reported dry eye symptoms beginning at day 28. No serious ocular or systemic adverse events were observed. No cases of intraocular inflammation, retinal vasculitis, endophthalmitis, or vision loss occurred during follow-up.

### Exploratory Functional and Anatomic Outcomes

All 9 treated subjects underwent BCVA assessment at each visit (Figure 2). The mean change in BCVA from screening to the OGX110 baseline visit, a period that included the antecedent anti-VEGF injection, was +1.4 ETDRS letters. Mean BCVA change from OGX110 baseline was +4.3 ETDRS letters at day 14 and +4.4 ETDRS letters at day 28. As the assessments were made on the same eye, this was subjected to a pair, one-sided t-Test, and found to be statistically significant at P < 0.05 at both time periods.a Six of 9 eyes gained at least 3 ETDRS letters within 4 weeks after OGX110 administration, whereas 1 eye experienced a decline in letter score (5 ETDRS letters). By day 56, the BCVA change was +2 ETDRS letters, with 6 participants maintaining at least a +4 ETDRS letter gain over baseline. There was no dose-dependency of the ETDRS letter gains. Given the small sample size, open-label design, and lack of a control group, these findings should be regarded as exploratory rather than confirmatory.

**Figure 2.**
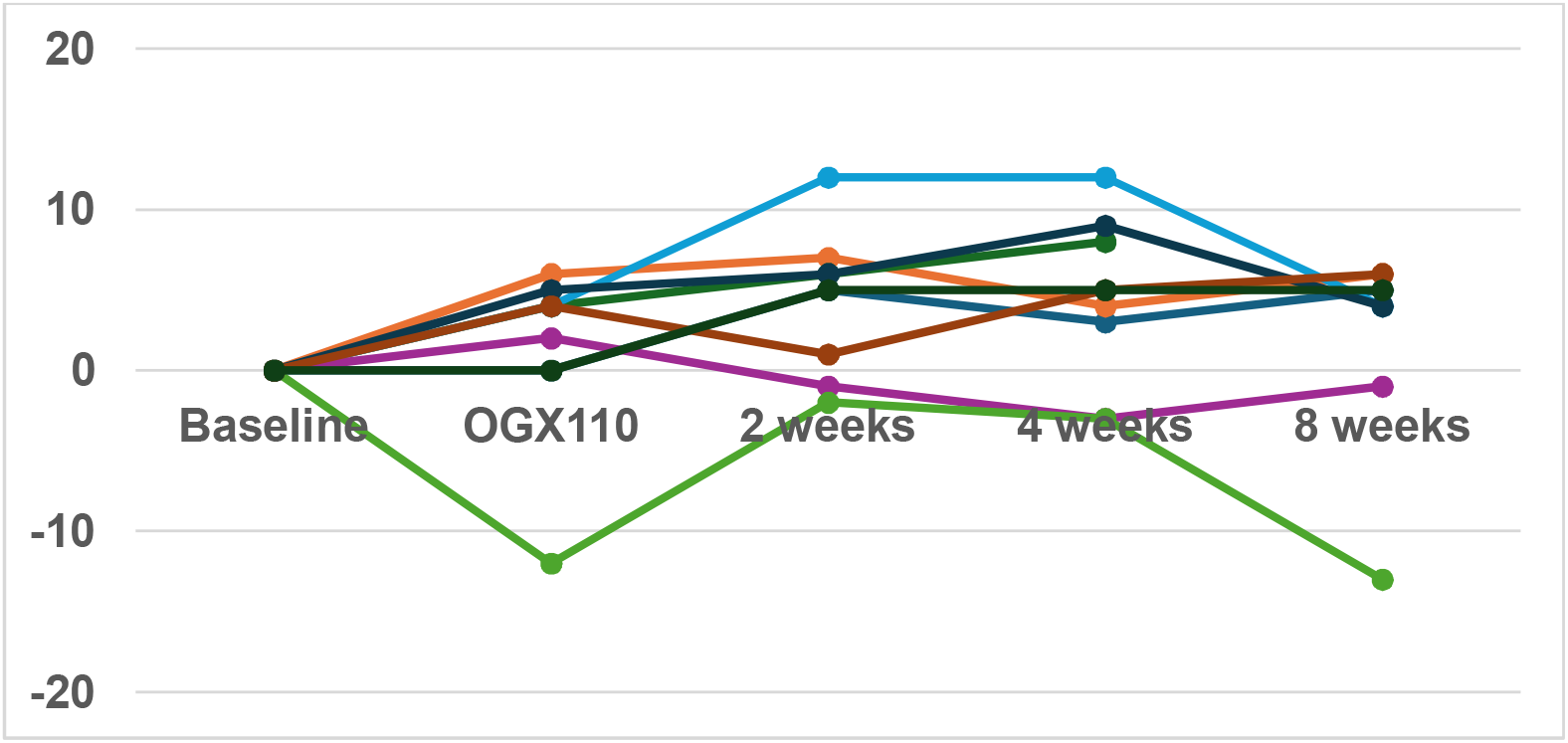
BCVA change in all 9 treated participants. The BCVA change from baseline reading within each participant (color-coded line) is noted for the two-month period after OGX110 IVT. The abscissa are the readings, not scaled to time, given the variance in the exact dates. The ordinate is ETDRS letters from the baseline reading.

Anatomic responses on SD-OCT were variable. Four of 9 eyes demonstrated a reduction in CRT after the antecedent anti-VEGF injection, and this reduction was generally maintained at 14 and 28 days after OGX110 administration. One additional eye showed a marked reduction in CRT after OGX110 despite minimal anatomic improvement after the preceding anti-VEGF injection (Figure 3). Mean CRT decreased by 44 um after the anti-VEGF injection and by 49 um at 14 days after OGX110 administration relative to screening values. Interpretation of these anatomic data is limited by heterogeneity in baseline fluid, disease chronicity, and anti-VEGF exposure.

**Figure 3.**
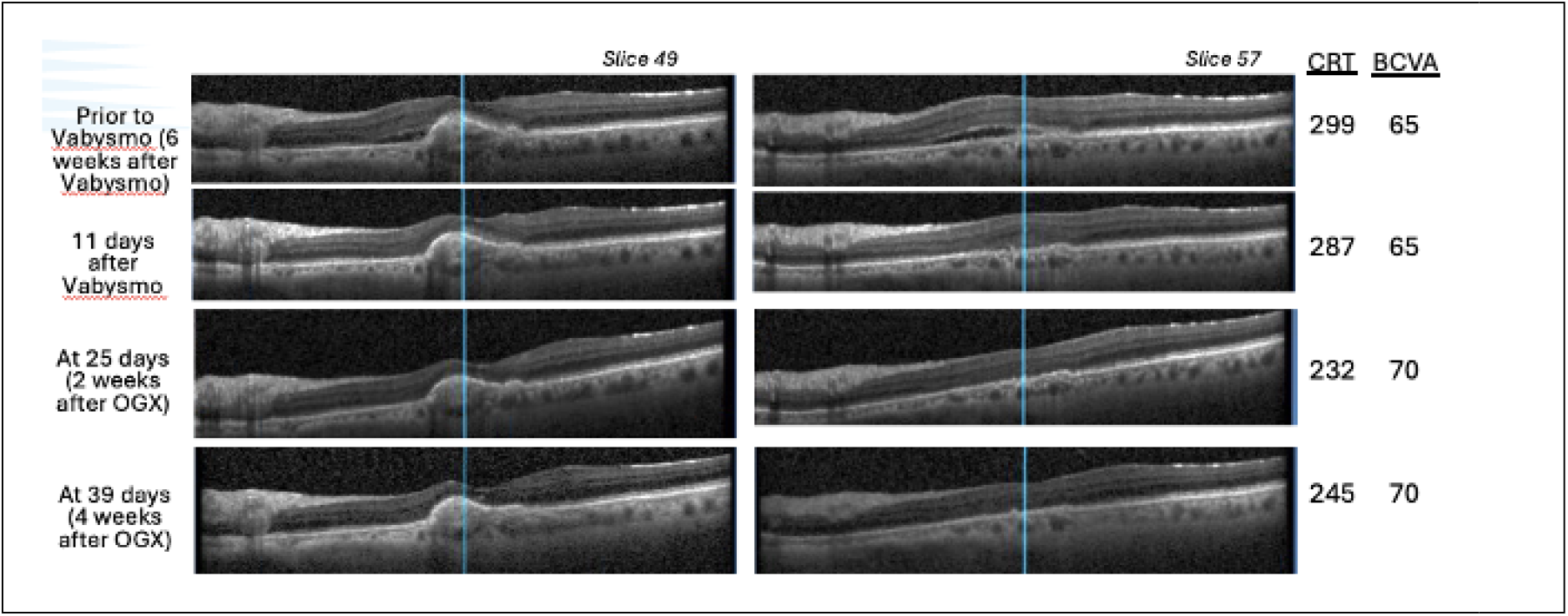
SD-OCT Images from Patient that shows augmented Response to OGX110 after anti-VEGF. Shown are two slices of the SD-OCT along with imager-generated CRT, and BCVA in ETDRS letters at screening, treatment and two follow-up visits for participant 9 (black line in Figure 2, above), who was treated with faricimab as part of routine clinical care, and then 11 days after received an OGX110 IVT as part of the study. Note that the faricimab had partial efficacy in diminishing subretinal fluid (SRF) in both slices but minimal impact on CRT or BCVA, which responded positively after the OGX110 IVT.

## Discussion

Herein, we communicate the first in human results of a new approach to treating neovascular age-related macular degeneration (nAMD). Treatment with OGX110, a peptide that activates the receptor CXCR3, pursues the hypothesis that nAMD represents a wound process that is chronically in the tissue replacement phase and does not progress to wound resolution. By providing pharmacologic levels of a ligand mimetic selective for CXCR3, the tissue can pharmacologically initiate the transition to wound resolution that includes involution of immature and pathological small blood vessels, reduction in matrix accumulation, recruitment and support of stem cells, and suppression of inflammation. While activation of CXCR3 has been shown to achieve near regenerative wound healing in various tissues in animal models, this is the first report of this pathway being therapeutically activated in humans and in the treatment of nAMD.

In this first-in-human phase I study, a single intravitreal injection of OGX110 was generally well tolerated in eyes with persistent nAMD receiving ongoing anti-VEGF therapy. No serious treatment-related adverse events were observed, and no dose-limiting toxicity emerged across the 3 dose levels studied. In addition, exploratory efficacy signals suggest possible improvements in visual acuity and anatomy when given in subjects with persistent disease activity despite previous and ongoing treatment with anti-VEGF agents. The initial exploratory findings provide for a benefit of the OGX110 IVT. The lack of dose effect was not unexpected as the doses were chosen not by highest tolerated, but based on expected binding affinity, with the mid-dose at calculated Kd and the higher and lower doses being only 2-fold different; thus all three doses should activate the CXCR3 receptors on the cognate cells.

However, these findings should be interpreted with caution. The study was small, uncontrolled, and open label, with substantial heterogeneity in disease duration, baseline anatomy, and prior anti-VEGF exposure. The short follow-up does not allow for evaluation of long-term effects. In addition, the temporal relationship between the antecedent anti-VEGF injection and OGX110 administration complicates attribution of short-term functional and anatomic changes to the investigational drug alone. Nonetheless, the functional and anatomic findings are hypothesis generating and support further study in a larger, controlled trial designed to better isolate treatment effect and characterize dose response.

Future studies should consider a randomized, controlled design, and more standardized eligibility criteria for persistent exudation. Additional doses and longer follow-up will also be important to assess durability, potential effects on fibrosis or atrophy, and whether repeated dosing is both safe and clinically meaningful.

In summary, OGX110 was generally well tolerated in this small phase I study of persistent nvAMD. Although exploratory outcome measures suggested possible adjunctive activity, definitive conclusions regarding efficacy cannot be drawn. These results of tolerability and potential efficacy support continued clinical development in appropriately controlled studies.

## Data Availability

All data produced in the present study are available upon reasonable request to the authors

## Acknowledgements

The trial and subsequent publication were sponsored by Ocugenix Therapeutics. No outside entities, nor AI, contributed to the writing of this publication.

---

**Supplementary Table 1. Inclusion Criteria**

**General Inclusion Criteria:**

1. Male or female subject 50 years of age or older
2. Subjects willing and able to provide a signed Informed Consent, as well as written local privacy requirements
3. Subjects willing and able to follow the study instructions and likely to complete all required study procedures and visits, as assessed by the investigator
4. Females of childbearing potential must consent to a pregnancy test before entering the study

**Ocular Inclusion Criteria in the Study Eye:**

1. Angiographically documented active CNV lesion (i.e., leakage on fluorescein angiography or subretinal, intraretinal, or sub-retinal pigment epithelium [sub-RPE] fluid on spectral domain optical coherence tomography [SD-OCT]) secondary to AMD, within the previous 10 weeks
2. CNV lesion size of up to 9-disc areas (22.5 mm^2^) with neovascularization composing 50% or more of the entire lesion area
3. Retinal hemorrhage less than 50% of total lesion area
4. Area of fibrosis within the CNV lesion must comprise less than 50% of the total lesion area
5. BCVA between 78 and 24 letters (approximate Snellen equivalent, 20/32 and 20/320), as assessed at the Screening Visit and confirmed at the Baseline Visit
6. Clear ocular media and adequate pupil dilation to permit good quality retinal imaging
7. Subjects must have received, 7 to 14 days prior to the Baseline Visit, an IVT injection in the study eye of an anti-VEGF ocular therapeutic approved for use in the United States

**Supplementary Table 2. Exclusion Criteria**

**General Exclusion Criteria:**

1. Female subjects of childbearing potential who are pregnant or unwilling to agree to practice at least one effective method of birth control for 6 months following administration of study medication.
2. Male subjects able to father a child who are unwilling to agree to practice an effective method of birth control (i.e., use of a condom) for 6 months following administration of study medication.
3. Subjects who, based on the investigator’s assessment, are too ill to likely complete the entire study
4. Subjects who, based on the investigator’s assessment, are not suitable for study participation
5. History or current evidence of severe hypersensitivity to latex or to any components of the study medication, or clinically relevant sensitivity to fluorescein dye
6. History or current evidence of a medical condition (systemic or ophthalmic disease, metabolic dysfunction, physical examination finding or clinical laboratory finding) that may, in the opinion of the investigator or medical monitor, preclude the safe administration of the study medication, adherence to the scheduled study visits, or safe participation in the study or affect the results of the study. Such conditions include but are not limited to unstable or progressive cardiovascular, cerebral vascular, pulmonary, Parkinson’s, liver or renal disease; depression, cancer, or dementia; evidence of prior vascular occlusion (e.g., stroke); and systemic or local hypercoagulable state.
7. Participation in any investigational study within 3 months prior to Visit 1 (Screening)
8. Subjects who have undergone major surgery within the previous 6 months (systemic or ocular) or who are likely to require major surgery within 6 months after Visit 1 (Screening)
9. Use of prohibited medications (list provided)

**Ocular Exclusion Criteria:**

1. History or evidence of the following surgeries/procedures in the study eye:
  a. Submacular surgery
  b. Vitrectomy
  c. Retinal detachment or retinal tear
  d. Incisional glaucoma surgery
2. Uncontrolled glaucoma or ocular hypertension, i.e., IOP ≥ 24 mmHg in the study eye
3. Cataract surgery or laser-assisted in situ keratomileusis (LASIK) performed within 3 months prior to Visit 1 (Screening) in the study eye
4. Anticipated need for cataract extraction in the study eye within 6 months of Visit 1 (Screening)
5. Ocular disorders in the study eye of a severity that could confound the interpretation of study results, compromise visual acuity, or require medical or surgical intervention during the study period. Such disorders include but are not limited to cataract, retinal vascular occlusion (current or prior), retinal detachment, retinal tear, macular hole, polypoidal choroidal neovascularization, geographic atrophy, CNV of any other cause (e.g., ocular histoplasmosis or pathological myopia), or autoimmune diseases of the eye or nervous system.
6. Current vitreous hemorrhage in the study eye
7. Spherical equivalent of the refractive error in the study eye demonstrating more than 8 diopters of myopia
8. History or evidence of penetrating ocular trauma in the study eye
9. Subject with congenital eye malformations in the study eye
10. Subject with uveitis or history of uveitis in either eye
11. Subject with any active ocular inflammation or ocular infection (i.e., bacterial, viral, parasitic, or fungal) in either eye
12. Periocular or ocular/intraocular infection or inflammation in either eye (e.g., infectious conjunctivitis, keratitis, scleritis, endophthalmitis) within 3 months prior to Visit 1 (Screening)
13. Aphakia in the study eye or break in the posterior capsule in the study eye, unless it is a small break resulted from a yttrium aluminum garnet (YAG) laser posterior capsulotomy in association with prior posterior intraocular lens implantation

**Supplementary Table 3:** Schedule of Visits and Procedures Following Single Escalating Doses of OCU-10-C-110 for Injection.

| Procedure | Screening | Day 0<br>Baseline | Day 1 | Week 2 | Week 4 | Exit Visit<br>Week 8 |
| --- | --- | --- | --- | --- | --- | --- |
| Visit Window (Days) | **-30 to -7**<br>**days** | **0 days** | **+ 1 day** | **± 4**<br>**days** | **± 5**<br>**days** | **± 5 days** |
| Visit Number | **1** | **2** | **3** | **4** | **5** | **6** |
| Informed Consent/Demographics<sup>a</sup> | **X** |  |  |  |  |  |
| Medical & Ophthalmic History | **X** |  |  |  |  |  |
| Concomitant Medications | **X** | **X** | **X** | **X** | **X** | **X** |
| Concurrent Procedures | **X** | **X** | **X** | **X** | **X** | **X** |
| Adverse Events | **X** | **X** | **X** | **X** | **X** | **X** |
| General Physical Exam<sup>b</sup> | **X** |  |  |  |  |  |
| Pregnancy Exam<sup>c</sup> | **X** |  |  |  |  | **X** |
| Clinical Laboratory Exams<br>(hematological and blood chemistry) | **X** |  |  |  |  | **X** |
| Best Corrected Visual Acuity (ETDRS) | **OU** | **SE** | **SE** | **SE** | **SE** | **OU** |
| Complete Ophthalmic Exam<sup>d</sup> | **OU** | **SE** | **SE** | **SE** | **SE** | **OU** |
| SD-OCT<sup>e</sup> | **OU** | **SE** |  | **SE** | **SE** | **OU** |
| Fluorescein Angiography | **OU** |  |  |  |  | **OU** |
| Dilated Fundus Photography | **OU** | **SE** |  |  |  | **OU** |
| Administration of OCU-10-C-110 for<br>Injection |  | **SE** |  |  |  |  |
| Administration of anti-VEGF<sup>f</sup> |  |  |  |  | **SE PRN** | **SE PRN** |
| Complete Exit Form |  |  |  |  |  | **X** |

Abbreviations: ETDRS = Early Treatment Diabetic Retinopathy Study, IOP = intraocular pressure, OU = both eyes, PRN = *pro re nata* (as needed), SD-OCT = spectral domain optical coherence tomography, SE = study eye, VEGF = vascular endothelial growth factor
- Demographic data includes height, weight, eye color and smoking history - General Physical Exam to include blood pressure and pulse rate - Females of childbearing potential only; additional pregnancy tests may be performed at any time/day during the study - Complete Ophthalmic Exam includes: external examination of the eye and adnexa, screening for eyelid/pupil responsiveness, slit-lamp biomicroscopy, indirect ophthalmoscopy, dilated fundus examination and IOP. IOP measurements will be taken prior to pupil dilation. If possible, IOP measurements should take place at approximately the same time of the day at each visit and with the same equipment - The same SD-OCT instrument should be used for an individual participant through the entire study - Anti-VEGF administration will be performed PRN (as needed) at the Week-4 and Week-8 Visits

